# Explaining the stable coexistence of drug-resistant and - susceptible pathogens: the Resistance Acquisition Purifying Selection model

**DOI:** 10.1101/2023.12.07.23299709

**Authors:** Pleuni S Pennings

## Abstract

Drug resistance is a problem in many pathogens. While overall, levels of resistance have risen in recent decades, there are many examples where after an initial rise, levels of resistance have stabilized. The stable coexistence of resistance and susceptibility has proven hard to explain – in most evolutionary models, either resistance or susceptibility ultimately “wins” and takes over the population. Here, we show that a simple model, mathematically akin to mutation-selection balance theory, can explain several key observations about drug resistance: (1) the stable coexistence of resistant and susceptible strains (2) at levels that depend on population-level drug usage and (3) with resistance often due to many different strains (resistance is present on many different genetic backgrounds). The model is applicable to resistance due to both mutations and horizontal gene transfer (HGT). It predicts that new resistant strains should continuously appear (through mutation or HGT and positive selection within treated hosts) and disappear (due to a fitness cost of resistance). The result is that while resistance is stable, *which* strains carry resistance is constantly changing. We used data from a longitudinal genomic study on *E. coli* in Norway to test this prediction for resistance to five different drugs and found that, consistent with the model, most resistant strains indeed disappear quickly after they appear in the dataset. Having a model that explains the dynamics of drug resistance will allow us to plan science-backed interventions to reduce the burden of drug resistance.

**Graphical abstract:** 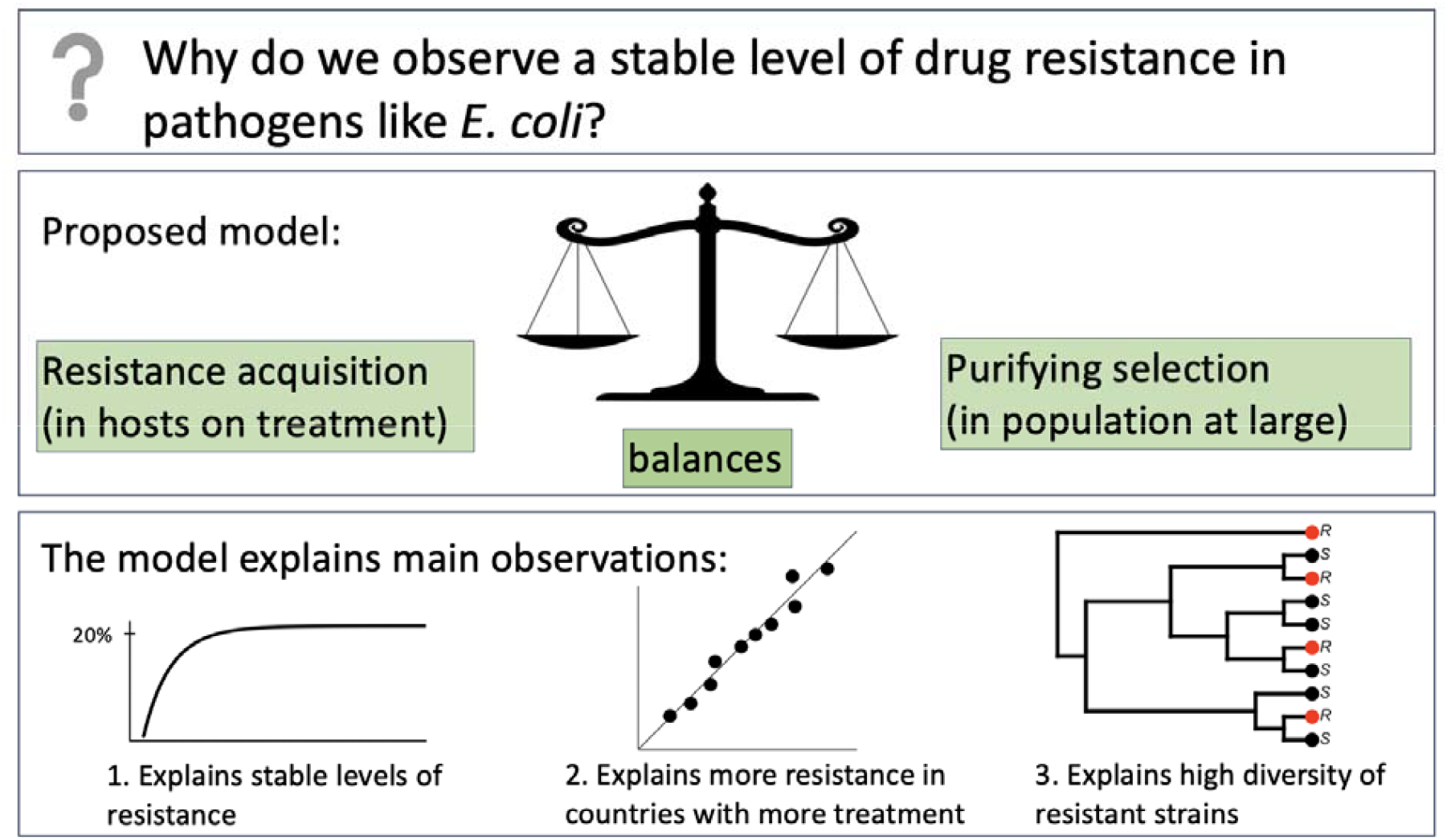

## Introduction

Drug resistance is common in viruses (e.g., HIV), bacteria (e.g., *S. aureus, E. coli, M. tuberculosis)*, parasites (e.g., *P. falciparum*) and fungi (e.g., *Candida* species), making it a major public health threat (Murray et al. 2022). What can we do to prevent the rise of drug-resistant strains? Or to revert the trend if the level of resistance is already high? To plan successful science-backed interventions, we must understand what determines the level of drug resistance in a given pathogen. Therefore, a need exists for better models to explain current resistance levels, predict how resistance rates will change over time and identify which interventions would most reduce the burden of resistance. Here, we present a simple model that does all these things.

Any model that might be used to predict drug resistance levels for different pathogens must be able to explain several observations that are currently not well explained (Davies et al. 2019; Krieger et al. 2020; Colijn et al. 2009). First, while overall drug resistance has increased over the years, in many cases, an initial increase in resistance is followed by stable coexistence between drug-resistant and drug-sensitive strains. For an example, see **Figure 1A** on quinolone resistance in *E. coli* in Europe. Other examples of stable coexistence of resistant and susceptible strains can be found in *S. pneumoniae* (Colijn et al. 2009; Krieger et al. 2020), *S. aureus* (Diekema et al. 2019) and HIV (Rhee et al. 2019; Rocheleau et al. 2018). Second, a positive linear relationship has been observed between treatment intensity at the population level and drug resistance levels (Goossens et al. 2005) (**Figure 1B)**. And third, many different resistant strains (i.e., resistance on different genetic backgrounds) often segregate at the same time in a pathogen population (Nübel et al. 2008; Rhee et al. 2019) (see **Figure 1C** for an example of resistance to the quinolone drug ciprofloxacin in *E. coli* in the UK; resistance appears repeatedly on the phylogenetic tree (van Nouhuijs et al. 2022; Kallonen et al. 2017).

**Figure 1.**
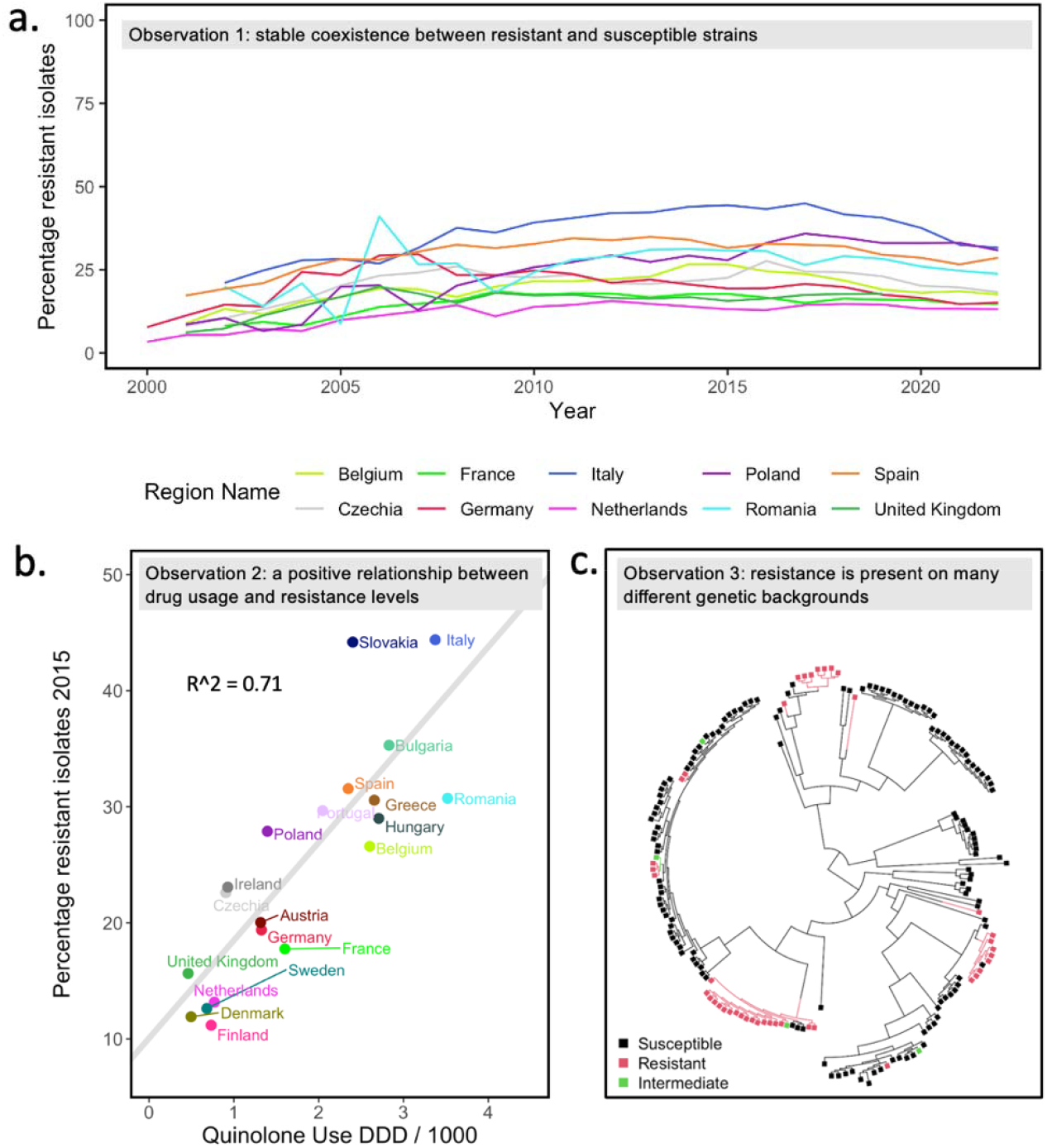
Three key observations about drug resistance, illustrated with data on resistance to quinolones in E. coli. a. Observation 1. Typically, an initial increase in resistance is followed by stable coexistence between drug-resistant and drug-sensitive strains. Each line depicts the fraction of E. coli strains resistant to quinolones in country over two decades in the 10 most populous countries (formerly) in the European Union (EU). **b**. Observation 2. Treatment intensity at the country level is correlated with drug resistance levels. A strong positive correlatio exists between quinolone usage and quinolone resistance levels in 20 countries in the EU. Usage is measured i “defined daily dose” per 1000 people. For ciprofloxacin, the defined daily dose is 1 gram. To go from DDD/1000 t rate of treatment per person, per year, we divide by 1000, multiply by 365 days, and divide by 5 under the assumption that 5 defined daily doses equal one course of antibiotics. Publicly available data from the Surveillance Atlas of Infectious Diseases were used to create Figures 1a and 1b (ECDC 2023; 2018). **c**. Observation 3. Many different resistant strains often segregate at the same time. Quinolone (specifically ciprofloxacin) resistance in bacteremia patients in the UK is due to many different origins of resistance (here E. coli phylogroup D and F are shown). Data from (Kallonen et al. 2017), figure reproduced from (van Nouhuijs et al. 2022).

The second observation feels intuitive to most of us: If we use more drugs, we get more drug resistance. But the first observation of stable coexistence, is puzzling: if there is selection *for* drug resistance, why doesn’t the level of resistance keep increasing until it reaches 100%? The third observation of resistance in many strains or genetic backgrounds, can be seen in many phylogenetic trees in articles about drug resistance, but usually does not receive a lot of attention. For example, in a large study of HIV isolates in Northern California, Rhee and colleagues found 82 phylogenetic clusters with one or more drug-resistance mutations (Rhee et al. 2019). In an *E. coli* dataset from the UK, 31 independent phylogenetic clusters with ciprofloxacin-resistant strains were found (van Nouhuijs et al. 2022), and Casali and colleagues found 106 independent phylogenetic clusters with pyrazinamide resistance in *M. tuberculosis* (Casali et al. 2014). These results show that resistance has evolved *de novo* or been acquired through horizontal gene transfer (HGT) many times (Nubel et al. 2008; Shin, Choi, and Ko 2012).

Several different explanations have been put forward to explain the coexistence of resistance and susceptibility and the positive relationship between treatment and resistance levels (Colijn et al. 2009; Krieger et al. 2020). These explanations have usually focused on mechanisms that require population structure or coinfection to create stabilizing selection (Colijn et al. 2009; Krieger et al. 2020; Cobey et al. 2017; Kouyos, Klein, and Grenfell 2013; Blanquart et al. 2018). We propose a simpler model akin to mutation-selection balance. Our model requires few assumptions, which means it could be applicable to a large variety of pathogens, and few parameters, which makes it easy to use.

We call our model the “resistance acquisition and purifying selection” (RAPS) model and will show that it can explain all three key observations about drug resistance listed previously. We will also describe which observations cannot directly be explained by the RAPS model. Note that when resistance is due to HGT, we don’t aim to explain why genetic elements that confer drug resistance exist or how they evolved, but rather why they are carried by a stable frequency of strains in pathogen populations.

### The resistance acquisition – purifying selection (RAPS) model

Here we show that the RAPS model based on classic mutation-selection balance can explain the thre striking observations of drug resistance at the population level: (1) a stable coexistence of resistance and susceptibility, (2) a positive relationship between drug usage and resistance levels, and (3) the simultaneous existence of many drug-resistant strains. Note that our goal here is not to determine whether the RAPS model can better explain the data than the models proposed by others. Instead, we provide a proof of concept to show that the model can, in principle, lead to the main observations and should be considered as a possibility. The RAPS model is applicable to resistance acquisition due to *de novo* mutation or HGT (though see discussion for limitations).

The RAPS model is based on the classic mutation-selection balance model, because this model has already been used to explain a stable level of deleterious mutations and the existence of multiple alleles that carry a deleterious mutation (Wakeley 2008; Hartl and Campbell 1982). Mutation-selection balance is a well-understood dynamic equilibrium, and we can borrow mathematical results from that theory directly here (Baum et al. 2017; Hartl and Clark 2006).

While mutation-selection balance has been considered by other authors in the context of drug resistance (Boni and Feldman 2005; Rodrigues, Gomes, and Rebelo 2007; Davies et al. 2021; Colijn et al. 2009), it has generally not received much attention as a potential explanation for intermediate drug resistance levels. Some authors mentioned mutation-selection balance but dismissed it as a viable model because they assumed that mutation-selection balance can only lead to resistance frequencies close to 0% or close to 100% (Colijn et al. 2009). Others argued that the rate of acquisition of resistance would not be high enough to explain observed frequencies (Davies et al. 2021). Yet others included mutation and selection as part of more complex models, but didn’t focus their analysis on it (Boni and Feldman 2005, Rodrigues, Gomes, and Rebelo 2007).

In the basic mutation-selection balance model, mutation creates deleterious alleles at rate μ per individual per generation and these alleles reduce the fitness of the individual carrying them by selection coefficient *s* per generation, see Table 1. Standard population genetic theory then predicts that the deleterious allele frequency is μ*/s* (Barton et al. 2007).

**Table 1.** Parameters for resistance acquisition-purifying selection (RAPS) model vs classical mutation-selection equilibrium model.

| Model parameter | Mutation – selection balance |  | Resistance acquisition – negative selection balance |  |
| --- | --- | --- | --- | --- |
| Generation of new variants | $\mu$ | mutation per generation | $t \cdot e$ | treatment and within-host evolution due to treatment per unit time |
| Selection against the variants | $s$ | deleterious selection coefficient per generation | $c$ | cost of drug resistance in the general population per unit time |
| Equilibrium frequency of the variant | $f = \frac{\mu}{s}$ | | $f = \frac{t \cdot e}{c}$ | |

To apply mutation-selection equilibrium theory to the problem of drug resistance, we make several adjustments. First, as others have done, we treat the population of a pathogen in a host as a single individual (Gordo et al. 2009). This means that a host can either be infected by a resistant pathogen or a susceptible pathogen and when evolution happens in a host it is instantaneous. The population size of the pathogen is assumed to be constant and controlled by an SIR model where hosts can be infected (I), susceptible to infection (S) or recovered (R).

Next, we assume that within-host populations evolve to become resistant at rate *t*·*e*, per unit of time. The treatment parameter *t* is the rate at which the pathogen encounters treatment. For example, if hosts have a probability of 5% per year to be treated with a given drug, *t* would be 0.05. The evolution parameter *e* is the probability (between 0 and 1) that the within-host pathogen population evolves to become resistant through mutation or the acquisition of a genetic element, given that the host is treated. Mathematically, *t*·*e* plays the role of the mutation parameter μ in the standard model (see Table 1). We’ll write the fraction of resistant strains as *f* and restrict its value to between 0 and 1. Only susceptible strains (*1-f* of the pathogen population) can evolve to become resistant.

We also assume that, except for within the treated host, the resistant type is less fit than the susceptible type. We refer to the difference in fitness as fitness cost *c*, which ranges between 0 and 1, so that its relative fitness of resistant strains is (*1-c*), when the fitness of the susceptible wildtype is set to *1* (though see Lenormand et al, 2018 for different use of the word cost related to resistance). The fitness cost makes that the resistant type less likely to be transmitted than the susceptible type (Wagner, Garcia-Lerma, and Blower 2012). Note that *c* needs to be measured in the same time unit as the rate of treatment (*t*). Mathematically, *c* plays the role of *s* (negative selection) in the standard mutation-selection model (see Table 1). The frequency of resistant strains will be reduced by the difference between the fitness of the resistant strains *(1-c)* and the average fitness of the pathogen population *(1-fc)*. For mathematical convenience, this term *((1-c) – (1-fc)*) can be rewritten as *-c(1-f)*.

Under this model, the frequency of resistant strains, *f*, will increase with *t*·*e*·*(1-f)* through positive selection within hosts and will be reduced by negative selection with *-c*·*(1-f)*· *f*. Thus, the change in frequency () can be written as:

An equilibrium will be reached when(, which means that the predicted equilibrium frequency of resistance will be either or, see **Figure 2**. This equilibrium result can explain the stability of the level of resistance (*f*), th linear relationship between treatment levels (through *t*) and the level of resistance (*f*), while allowing for evolution of new resistant strains to occur. For an alternative derivation of the same result starting from an epidemiological model, see methods.

**Figure 2:**
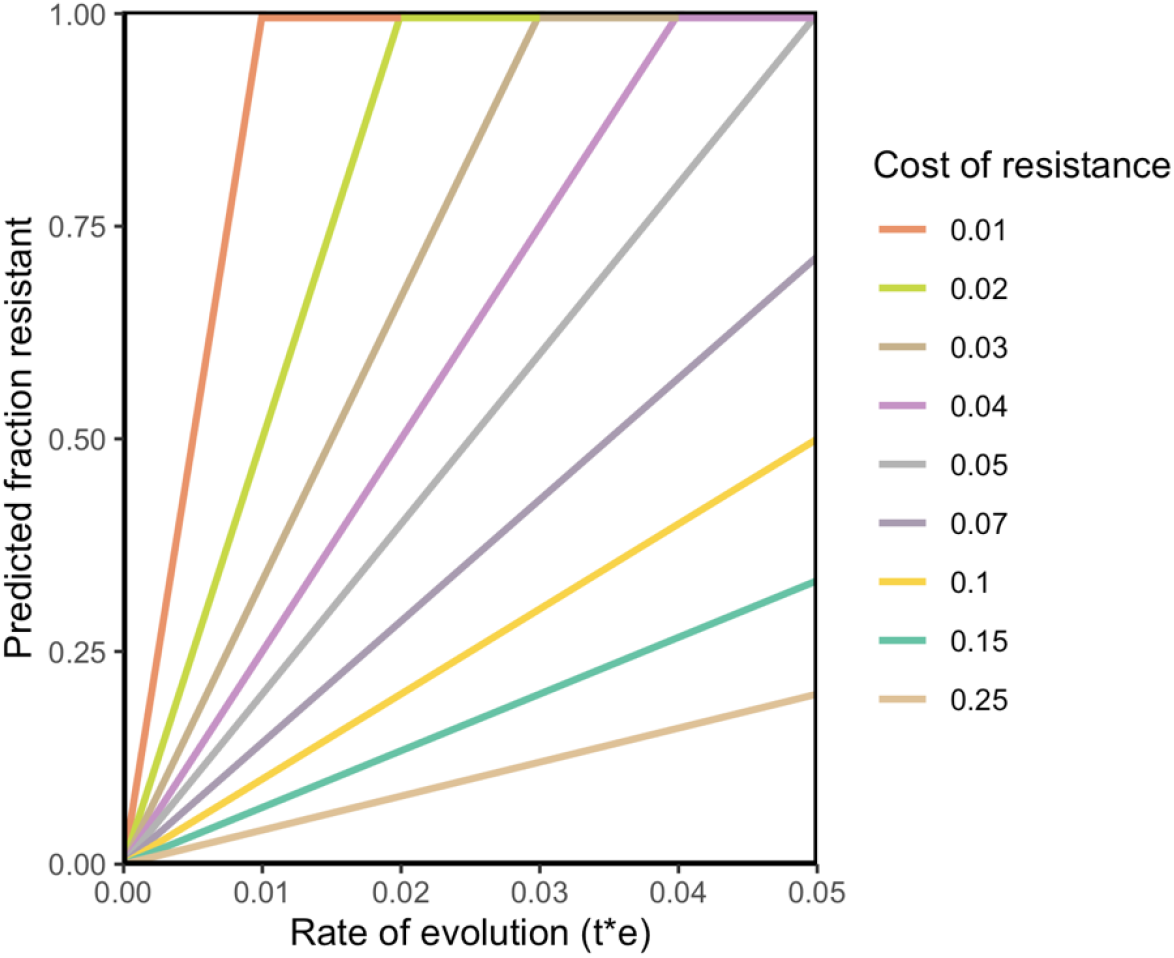
The level of resistance predicted by the Resistance Acquisition – Purifying Selection (RAPS) model for a population of pathogens is a function of the probability of resistance evolution per infection, t·e, and the cost of resistance, c. Higher levels of treatment lead to higher values of f, consistent with Key Observation 2 about drug resistance. Strains acquire resistance b mutation or through HGT. Resistant strains can be transmitted to other hosts, but with a lower rate of transmission compared to susceptible strains. c measures how much less fit resistant strains ar compared to susceptible strains.

### Genomic and phenotypic data

Using 16 years of publicly available data for resistance to several drugs in *E. coli* in Norway (Gladstone et al. 2021), we show that resistance acquisition is common and that most resistant strains are short-lived, which is consistent with the RAPS model. We used a dataset of *E. coli* genomes from bloodstream infections in Norway consisting of 3254 isolates from a surveillance program from 2002 - 2017. **Figure 3A** shows the percentage strains resistant to ampicillin, cefotaxime, ciprofloxacin, gentamicin and piperacillin-tazobactam in the dataset over time.

**Figure 3:**
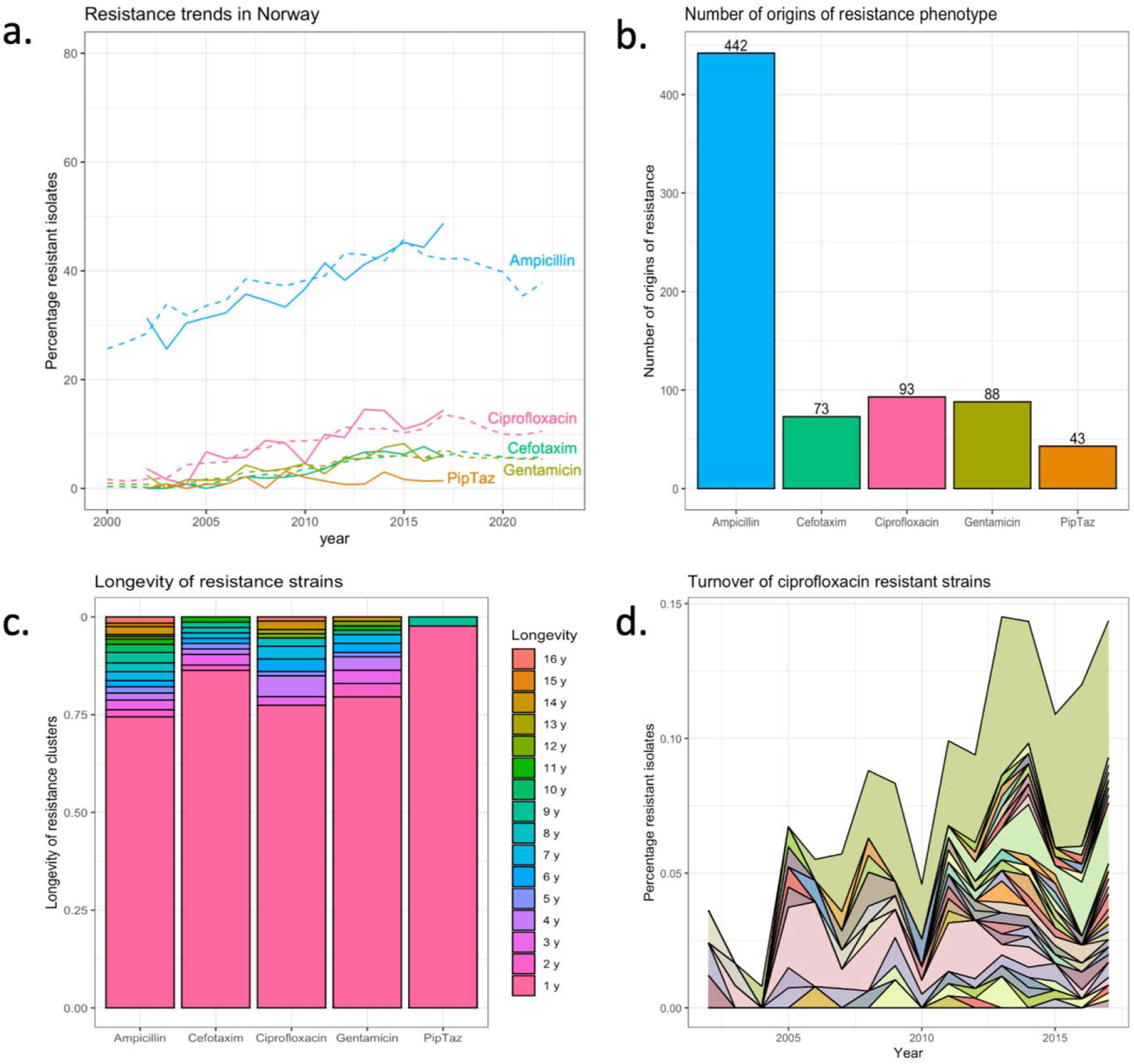
Dynamics of drug-resistant strains in Norway. PipTaz is piperacillin-tazobactam. **a**. Resistance trends in Norway for five drugs based on (ECDC 2023; Gladstone et al. 2021). **b**. Estimated number of origins of resistance to five drugs. **c**. Lifespans of resistant strains. **d**. Turnover of ciprofloxacin (quinolone) resistant strains. Each color represents a different resistant strain. The total of all resistant strains is the percentage ciprofloxacin-resistant strains as shown in panel **a**.

Using the phylogenetic tree and the phenotypic resistance data provided by the authors of the original paper (Gladstone et al. 2021), we estimated that ampicillin resistance evolved over 400 times on the tree, and resistance to the other drugs (cefotaxime, ciprofloxacin, gentamicin and piperacillin-tazobactam) evolved between 43 and 93 times (**Figure 3B**) [see Methods]. Note that the big difference in the number of evolutionary origins between ampicillin and the other drugs is consistent with the fact that ampicillin is an antibiotic that is much more often prescribed than the others (ECDC 2023).

We estimated the lifespan of individual resistant strains by determining, for each origin of resistance, in which years the resulting resistant strain was observed in the dataset. We found that for the five drugs we looked at, at least 75% of the strains were observed in only one year in the dataset (**Figure 3C**). We acknowledge that these lifespan estimates depend on the sampling density and are likely underestimates.

Next, we created an area plot that shows how most ciprofloxacin (quinolone) resistant strains are only seen for a short time span and have low frequencies, but that also shows a few longer-lived strains and one resistant strain that is long-lived and reaches a high frequency (**Figure 3D**). While the many strains that appear and disappear again are consistent with the RAPS model, the model cannot explain the behavior of the strain that increases in frequency over several years. In fact, it is unlikely that this increase is driven by the resistance to ciprofloxacin, instead, as others have suggested, it could be that by chance the ciprofloxacin resistance mutations landed on a background with positively selected elements and are hitchhiking to high frequency (Kallonen et al. 2017).

In summary, our analysis of the Norwegian dataset shows that resistant strains to five different drugs evolve often, yet most resistant strains do not reach high frequency and do not stay in the population for long, which is consistent with ongoing selection against resistance and with the RAPS model. It should be noted though that the number of different resistant strains that is observed cannot directly be interpreted in terms of model parameters, so a quantitative test of the model is not possible here. In addition, there are a few long-lived resistance strains which are unexplained by the RAPS model and which account for a significant portion of resistance in *E. coli*.

## Discussion

To understand, predict and influence levels of drug resistance in pathogens, we need mathematical models. Here we show that a simple model with strong positive selection for drug resistance within treated hosts and mild negative (purifying) selection in the rest of the population can explain three key observations about drug resistance in a variety of pathogens (**Figure 1**). First, the resistance acquisition – purifying selection (RAPS) model explains the stable coexistence of resistance and susceptibility in pathogen populations. Second, it explains the positive, linear relationship between the amount of drugs used and the frequency of drug resistance (**Figure 2**). And finally, it recapitulates the simultaneous existence of many different strains that carry a resistance mutation or genetic element at any one time (**Figure 1C**).

A major strength of the RAPS model is its simplicity. It relies on only a few assumptions and doesn’t require consideration of co-infection or spatial structure of the host population. One benefit of this simplicity is that the model is more likely to be applicable to different pathogen systems. The most important assumption we make is that resistance comes with a fitness cost for the pathogen, reducing its R_0. If that is not the case, resistant strains will outcompete susceptible strains (Cohen and Murray 2004). A major difference between our model and some previous models that attempt to explain stable coexistence of drug resistance and susceptibility (Krieger et al. 2020; Colijn et al. 2009) is that the RAPS model explicitly includes the creation and dying out of resistant strains. This can happen even when the specific genetic elements that confer resistance are stably present in the population. One reason why researchers may have chosen to ignore the creation and dying out of resistant strains is because it is assumed that drug-resistant strains are stable. However, stability at the phenotypic level doesn’t necessarily mean stability at the genetic level. New strains may evolve resistance in the same way as previous strains, even acquiring genetic elements from previous strains. The result is that the genetic element or the mutation may be stably present, but “who” carries it may always be changing (**Figure 3D**).

Another strength of our model is that it makes several testable predictions. 1). It predicts that for a given drug, the higher levels of resistance in countries with high drug usage is because resistance acquisition happens more often (and not because resistant strains are fitter). The best way to test this may be to compare the same country at higher and lower treatment levels. We expect that a doubling (halving) of the treatment level should lead to a doubling (halving) of the number of resistant strains in a sample of the same size. An interesting possible future study would therefore be to compare genomic data from a single country at different time points, if enough sequences are available. For example, in Germany in 2011 the fraction with ciprofloxacin resistance was 24%, 11 years later, in 2022, the resistance fraction was 15%. If the RAPS model is correct, we’d expect to see more origins of resistance in 2011 than in 2022.

2). Another testable prediction is that the lifespan of resistant strains in a population should depend on the cost of the resistance for the pathogen and not the level of treatment. Resistance strains in two different countries or at two different levels of treatment, should therefore have similar life spans if the RAPS model is correct. 3). In addition, the time scale at which resistance levels change when treatment habits change should depend on the cost of resistance. High costs mean short life spans, which means that a new equilibrium can be reached faster, and interventions should pay off more quickly. Intervention studies where ciprofloxacin (quinolone) use was reduced seem to indicate that this can have an effect on resistance levels in a few months (Sarma et al. 2015; Gottesman et al. 2009). Population genetic analysis suggests that costs of PipTaz resistance and Cefuroxime resistance are higher (van Nouhuijs et al. 2022), which should lead to shorter lifespans (see **figure 3C** for PipTaz) and faster effects of reduced usage.

Thanks to publicly available data, we were able to show that stability at the population level indeed hides dynamic changes at lower levels (see **Figure 3A-D**). Although the resistance phenotype is present at a stable level (**Figure 3A**), resistance is due to many evolutionary origins (**Figure 3B**) and 75% of resistant strains are only present for a year (**Figure 3C-D**). It will be of interest to see if more granular data (e.g., monthly surveillance from a hospital) would show more clearly the waxing and waning of resistant strains and would allow us to estimate the fitness cost of resistance which is not possible with the current data. A major open question is why there are some long-lived resistance strains while most are so short-lived. Are these long-lived strains hitchhiking with other genetic elements as others have suggested (Kallonen et al. 2017)? Do these strains occasionally lose the resistant phenotype, as would be expected if the resistance is a costly hitchhiker?

It is of interest to see if what we know about treatment levels and drug resistance evolution probabilities roughly fits with the RAPS model. Because we don’t know the typical time of an *E. coli* infection, we will use year as the unit of time here. We take the European data and estimate the slope of a linear model that goes through the origin (assuming no resistance if there is no treatment in a country), we find that the frequency of resistance goes up by about 10% for each 7% probability of treatment per year per person (assuming ciprofloxacin treatment is on average 5 days). Under the RAPS model, this means that the ratio between *e* (the probability of resistance evolution) and *c* (the fitness cost per year of resistance) should be 0.1/0.07 (because at equilibrium *f/t* = *e/c*). However, neither *e* nor *c* are well known from the literature. Yaffe et al (2023) studied evolution of resistance to the quinolone drug ciprofloxacin in the gut microbiome of healthy volunteers and estimated the probability of evolution of resistance to be around 10%, though when they only focused on the *E. coli* populations, they found that none of the 7 ciprofloxacin-susceptible *E. coli* populations in their study acquired resistance. Other studies with healthy volunteers or patients found that replacement by a resistant strain is more likely than evolution of ciprofloxacin resistance (de Lastours et al., 2012, Stracy et al. 2022). If *e* is 10% (as suggested by Yaffe et al. (2023)) then *c* must be around 7% per year if the RAPS model is correct.

We don’t know much about the fitness cost of resistance outside of the lab. The best way to determine if there is a cost of resistance in the absence of treatment is to stop all treatment with a given drug in a large area and see if and how fast resistance levels go down. In fact, in Israel, for a short time there was a policy to not use any ciprofloxacin in the country to preserve the drug for use in postexposure prophylaxis in case of anthrax bioterrorism. While the policy wasn’t followed perfectly, ciprofloxacin usage did go down for several months. Gottesman et al. (2009) observed a decrease in resistance from 12 to 9% in 6 months. Though the authors didn’t attempt to estimate the strength of selection against resistant strains, this strong reduction would suggest a *c* would need to around 50% per year or higher.

Though not as drastically as what happened in Israel, many European countries reduced the usage of quinolones in the 2010s. Data from the ECDC allow us to compare quinolone usage and ciprofloxacin resistance from 2011 to 2022 for the 20 largest European countries (**Figure 4)**. If the RAPS model is correct, resistance levels should go down when treatment levels go down. Indeed, in 15 of 16 countries where usage went down between 2011 and 2022, resistance also went down (**4A**). In addition, in 3 of 3 countries where usage went up, resistance went up (**4B**). In the UK, usage was stable, and resistance stayed at the same level as well. (**4C**). Finally, in only one country (Sweden) something unexpected happened: usage levels went down, but resistance levels went up (**4C**). Data in 19 out of 20 countries are therefore consistent with the RAPS model. These data support the idea that usage levels directly influence resistance levels and that there is a fitness cost of resistance which leads to a decline in resistance when usage goes down.

**Figure 4:**
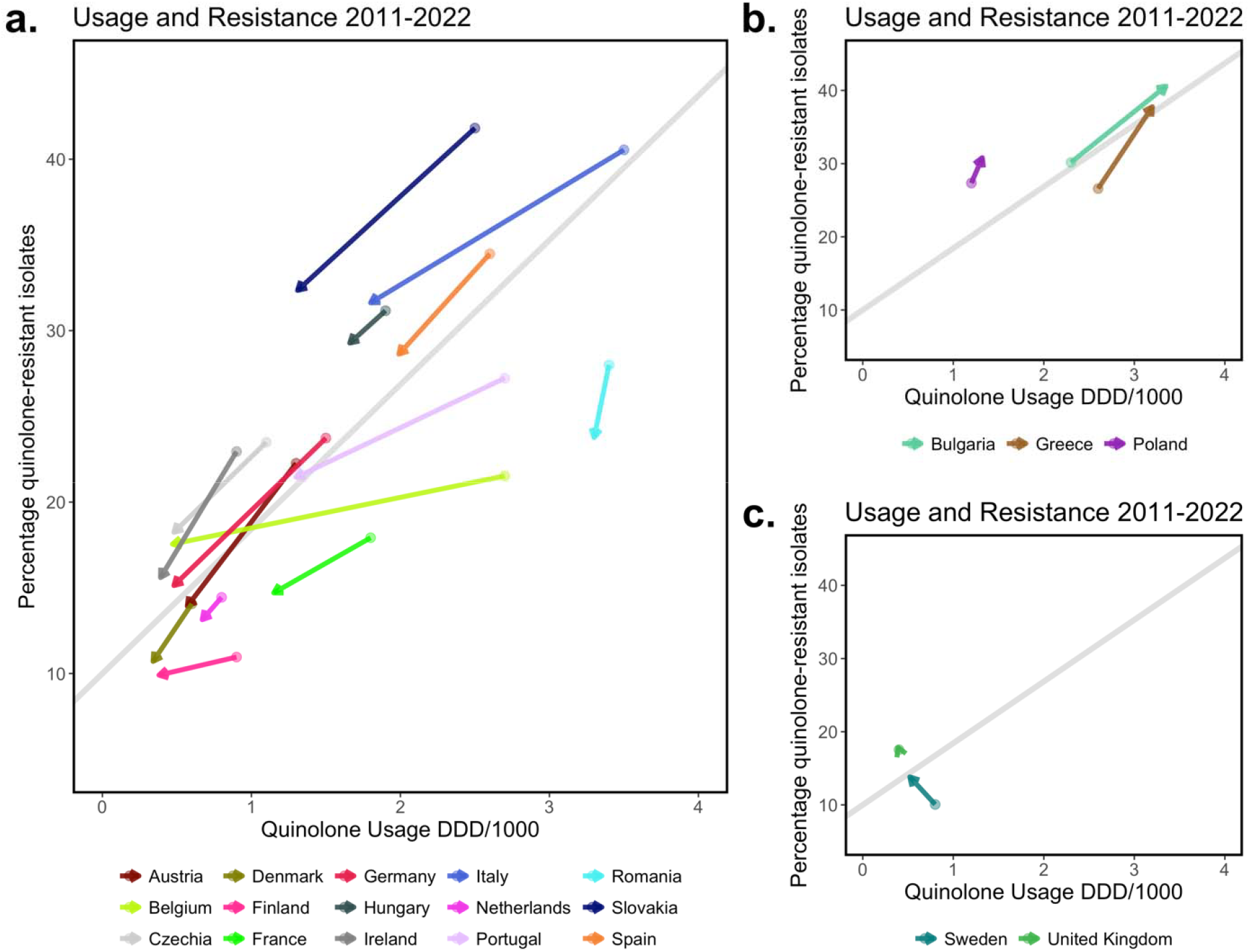
Changes in quinolone usage and quinolone resistance in the 20 most populous European countries (including the UK). a. In 15 countries, the level of usage of quinolones went down between 2011 and 2022 and the level of resistance to quinolones also went down during that time span. b. In thre countries, the level of usage of quinolones went up and the level of resistance to quinolones also went up. c. For the United Kingdom we only have data until 2019, but there was no change in either quinolon usage or quinolone resistance. Sweden is the only country where the usage and resistance moved in opposite directions. The level of usage of quinolones went down between 2011 and 2022, but the level of resistance to quinolones went up.

In conclusion, for ciprofloxacin (quinolone) resistance, we don’t know where new resistant strains are coming from, but studies suggest that *e* is on the order of 10% or lower (Yaffe et al 2023, de Lastours et al., 2012, Stracy et al. 2022). Only one study provides weak evidence for a strong cost (Gottesman et al 2009). However, ECDC data provides strong evidence that the fraction resistant strains in a population goes down when treatment levels are reduced over several years most likely because of a fitness cost (**Figure 4A**).

Our study has several limitations. One limitation is that the model considers only resistance to a single drug and not interactions between drugs or resistances. For example, when plasmids carry multiple resistance genes, resistance to one drug can be acquired due to selection for resistance to another drug. We also assumed that treatment does not affect transmission. If treatment itself is associated with strongly reduced transmission (as is the case in HIV), then it would be useful if the model included the treatment state of hosts, where the transmission probability of hosts on treatment is significantly lower than those not on treatment (Kühnert et al. 2018; Stadler et al. 2013). The cost of resistance may vary between strains and may be affected by compensatory mutations which we have not included (Cohen and Murray 2004; Hinz et al. 2023). While the RAPS model can be applied to resistance due to mutations or horizontally transferred genetic elements, it assumes a constant rate at which resistance is acquired, which means that for it to be applicable to resistance due to HGT, we assume that there is a reservoir of resistance genes outside the modeled species. In a model where resistance genes are acquired at a higher rate when they are common, the relationship between usage and resistance would be accelerating and not linear.

If stable levels of resistance mask the ongoing appearance of new resistant strains, this may have implications for machine learning methods that are used to predict drug resistance phenotypes from genomic data. At any given year and location, a machine learning algorithm may learn to recognize certain strains responsible for much of the resistance burden in a species. Yet, over time, these resistant strains may be replaced by others, which means that we likely need to re-train such models for current and local situations (Moradigaravand et al. 2018, Orcales, Moctezuma Tan et al. 2024).

In conclusion, we propose a simple model that can explain why drug resistance can stably co-exist with susceptible strains at levels that depend on drug usage. The model also explains why drug resistance is often present in many different strains (phylogenetic clusters, genetic backgrounds or sequence types) and predicts that resistant strains evolve and disappear again regularly. Data for resistance to several drugs in *E. coli* in Norway and data on usage and resistance changes over time in 20 European countries largely consistent with predictions. On the other hand, the model does not explain why some resistant strains are seen over many years (**Figure 3D**). In addition, data on emergence of quinolone-resistant strains (Yaffe et al 2023, de Lastours et al., 2012, Stracy et al. 2022) and data on the cost of quinolone resistance (Gottesman et al 2009) are not consistent with the RAPS model. In future studies different elements of the RAPS model and elements of other models could be combined to gain a more complete understanding of the dynamics of drug resistance.

The RAPS model has the potential to help us understand, predict and influence drug resistance levels and because of its simplicity, its assumptions and predictions can be readily tested in many different pathogen systems such as *S. pneumoniae* (Colijn et al. 2009; Krieger et al. 2020), *S. aureus* (Diekema et al. 2019) and HIV (Rhee et al. 2019; Rocheleau et al. 2018)).

## Data Availability

All data and code are on github: https://github.com/pleunipennings/CoexistencePaper

## Acknowledgements

I am grateful for comments from and discussions with Kristin Harper, Joachim Hermisson, Sarah Cobey, Alison Feder, Nandita Garud, Oana Carja, Rasmus Nielsen, Alison Hill, Scott Roy, Brandon Ogbunu, Ruth Hershberg, several anonymous reviewers, and the students in the SFSU CodeLab. Olivia Pham helped with figure 1.

## Methods

All code and data are available on Github: https://github.com/pleunipennings/CoexistencePaper.

Figure 1A European surveillance data

Quinolone resistance over time in 10 European countries.

Data were downloaded from the European Surveillance Atlas for Infectious Diseases. Link https://atlas.ecdc.europa.eu/public/index.aspx.

As health topic, we chose “Antimicrobial resistance”, as subpopulation “*Escherichia coli*|” and “fluoroquinolones” and as indicator “percentage resistance”. We then downloaded a csv file with data for all available years and regions (named ECDC_surveillance_data_Antimicrobial_resistance_complete_DownloadApril2024.csv in the Github repository). For Figure 1A we included the 10 most populous countries in the EU, including the United Kingdom: Germany, United Kingdom, France, Italy, Spain, Poland, Romania, Netherlands, Belgium, Czech Republic (ECDC 2023).

Figure 1B European surveillance data

For figure 1B, we used the percentage fluoroquinolone resistance in 2015 from the dataset described for Figure 1A, but now for the 20 most populous countries. This included in addition to the first 10: Sweden, Portugal, Greece, Hungary, Austria, Bulgaria, Denmark, Finland, Slovakia, Ireland.

For data on the use of fluoroquinolones in these countries, we downloaded data from https://www.ecdc.europa.eu/en/publications-data/antimicrobial-consumption-annual-epidemiological-report-2015 which included “Downloadable tables: Antimicrobial consumption - Annual Epidemiological Report for 2015” (ECDC 2018). The file is named 2015_data_Table_D6_J01M_quinolone antibacterials_trend_community_sparklines.xlsx on Github.

Figure 1C *E. coli* phylogenetic tree Kallonen dataset from Nouhuijs et al.

The phylogenetic tree in Figure 1C was reproduced from (van Nouhuijs et al. 2022). Data presented here include only *E. coli* strains from phylogroup D and F. The genomes and phenotypes were described in (Kallonen et al. 2017). Briefly, a core genome alignment was used to create a phylogenetic tree, then we simulated a single history of the resistance phenotype using the “simmap” function in Phytools (Bollback 2006; Revell 2012), the simulated history is shown on the tree with resistant branches in red, susceptible branches in green and intermediate branches in green.

Figure 3A Resistance trends in Norway for five drugs

Data are from Gladstone, 2021 (Gladstone et al. 2021) and from the ECDC Surveillance Atlas (ECDC 2023).

Figure 3B Estimated number of origins of resistance to five drugs

We used the tree and metadata provided by Gladstone, 2021 (Gladstone et al. 2021). Intermediate and susceptible strains were lumped in one category. We simulated a single history of the resistance phenotype using the “simmap” stochastic character mapping function in Phytools (Revell 2012) which is based on methods by (Bollback 2006; Huelsenbeck, Nielsen, and Bollback 2003). Next, we used a custom R script to identify the last resistant ancestor for each resistant tip of the tree. If two tips share a last resistant ancestor, we assume that they are resistant due to the same evolutionary origin of resistance. This evolutionary origin may be due to a mutation, several mutations or horizontal gene transfer – our approach is based on the phenotypes, not the genotypes. Based on the simmap results, we count the number of times resistance was acquired on the tree.

**Supplementary figure 1.**
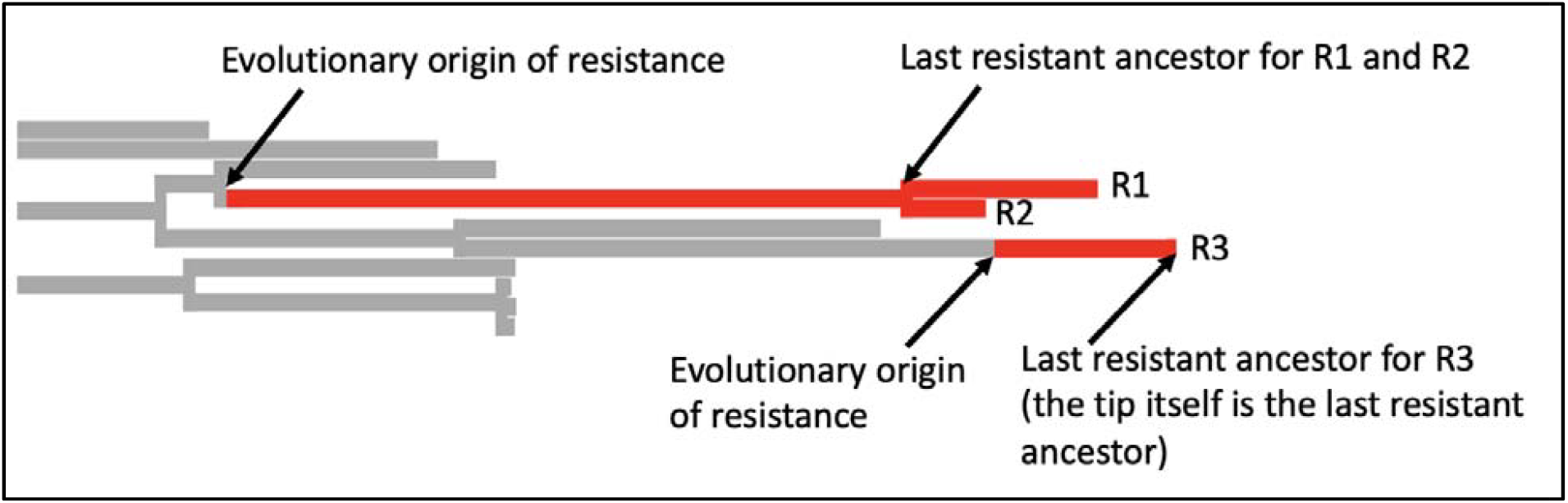
In this example, there are three resistant tips. The branches are colored according to the simmap stochastic character mapping. R1 and R2 share a last resistant ancestor and are therefor assumed to be resistant due to the same evolutionary event in their history, we consider them part of th same resistant cluster or resistant strain. For R3, the last resistant ancestor is itself. It is the only member of the resistant cluster or strain.

Figure 3C Lifespans of resistant strains

Using the results from analysis described for 3B and the metadata from (Gladstone et al. 2021) to determine for each inferred resistance cluster for how many years it was observed in the data. For example, if R1 was a sample from 2005 and R2 was a sample from 2007, then we infer that the lifespan of this cluster was 3 years.

Figure 3D Turnover of ciprofloxacin (quinolone) resistant strains

Using the results from analysis described for 3B and the metadata from (Gladstone et al. 2021) to determine for each inferred ciprofloxacin resistance cluster what its frequency was in each year.

Figure 4

Usage data were combined from the data we used for Figure 1B and a later ECDC report antimicrobial-resistance-ESAC-Net-report-downloadable-tables-2022.xls downloaded from the website: https://www.ecdc.europa.eu/en/publications-data/downloadable-tables-antimicrobial-consumption-annual-epidemiological-report-2022.

Drug resistance data were from the same source as for Figure 1A.

### Alternative derivation of mathematical result

The equilibrium level of resistance *t*·*e/c* can be derived starting from an epidemiological model. Dynamics of drug-sensitive strains *I*:

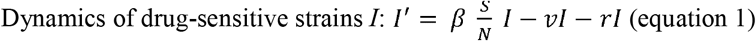

Dynamics of drug-resistant strains

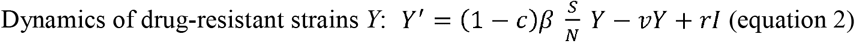

Here *c* is the transmission fitness cost, *β* is the transmission rate, *S/N* is the fraction of the population susceptible to infection, *v* is the recovery rate (which can include background mortality) and *r* is the rate at which individuals carrying a sensitive strain are treated and evolve drug-resistance. The probability that an individual carrying a drug-sensitive strain leaves from that class because they develop drug-resistance (rather than recovering) is given by *r/(r+v)*, and this is equivalent to (*t·e*).

We can solve for equilibrium fraction of susceptible individuals using equation 1 by setting *I*′ to 0 and solving for (*S/N*): *S/N* at equilibrium = (*v +r*)/*β*.

(Note that if the fraction (*v +r*)/*β*is larger than 1, the disease cannot spread in the population).

We can now substitute the equilibrium value of S/N into equation 2: 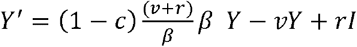

If we set *Y*′ to 0, we find 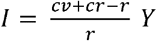, which we can use to calculate the fraction of resistant strains in the population at equilibrium is 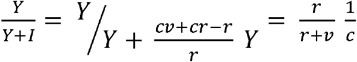.

Because 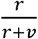 is equivalent to (*t*·*e*), we find that in this model too, the fraction resistant strains at equilibrium will be (*t*·*e*) / *c*.

